# Viral transcript and tumor immune microenvironment-based transcriptomic profiling of HPV-associated head and neck squamous cell carcinoma identifies subtypes associated with prognosis

**DOI:** 10.1101/2024.09.16.24313563

**Authors:** Anastasia Nikitina, Daria Kiriy, Andrey Tyshevich, Dmitry Tychinin, Zoia Antysheva, Anastasia Sobol, Vladimir Kushnarev, Nara Shin, Jessica H. Brown, James Lewis, Krystle A. Lang Kuhs, Robert Ferris, Lori Wirth, Nikita Kotlov, Daniel L. Faden

## Abstract

Human papillomavirus (HPV)-associated head and neck squamous cell carcinoma (HPV-positive HNSCC) has distinct biological characteristics from HPV-negative HNSCC. Using an AI-based analytical platform on meta cohorts, we profiled expression patterns of viral transcripts and HPV viral genome integration, and classified the tumor microenvironment (TME). Unsupervised clustering analysis revealed 5 distinct and novel TME subtypes across patients (immune-enriched, highly immune and B-cell enriched, fibrotic, immune-desert, and immune-enriched luminal). These TME subtypes were highly correlated with patient prognosis. In order to understand specific factors associated with prognosis, we used unsupervised clustering of a HPV-positive HNSCC cohort from The Cancer Genome Atlas (TCGA) (n = 53) based on HPV transcript expression, and identified 4 HPV-related subtypes (E2/E5, E6/E7, E1/E4 and L1/L2). Utilizing both viral transcript and TME subtypes, we found that the E2/E5 HPV subtype was associated with an immune-enriched TME and had a higher overall survival compared to other subtypes. The E2/E5 subtype was also enriched for samples without HPV-genome integration, suggesting that episomal HPV status and E2/E5 expression pattern may be associated with an inflamed microenvironment and improved prognosis. In contrast, E6/E7 subtype samples were associated with the fibrotic and immune-desert TME subtypes, with lower values of T-cell and B-cell gene expression signatures and lower overall survival. Both E1/E4 and L1/L2 subtypes were associated with the immune-enriched luminal subtype. Our results suggest that HPV- transcript expression patterns may drive modulation of the TME and thereby impact prognosis.

## Introduction

Human papillomavirus (HPV)-associated head and neck squamous cell carcinoma (HPV- positive HNSCC) is the most common HPV-associated malignancy in the United States. Currently, there is a lack of predictive biomarkers, which has stagnated efforts to personalize treatments. Thus, at present, many patients receive excessive treatment with severe lifelong side effects, while others are undertreated and at risk for relapse. Here, we describe the use of a next generation sequencing (NGS) data-based analytical platform to characterize the expression patterns of viral transcripts, the tumor microenvironment (TME), and viral genome integration and associate these features with overall survival (OS).

Growing evidence has indicated that the TME, a complex milieu of various cell types, including immune cells, fibroblasts, and endothelial cells, plays a significant role in a tumor’s growth and progression, and modulates its response to therapy [1–4]. Given the immunogenic nature of HPV, it is hypothesized that the TME in HPV-positive HNSCCs would exhibit distinct characteristics compared to HPV-negative HNSCC [5]. Furthermore, the expression patterns of viral transcripts within the tumor could play a role in modulating the TME and subsequently, the tumor’s behavior and prognosis [6].

In this study, we aimed to dissect the TME in HPV-positive HNSCCs using functional gene expression signatures (Fges) and explore the associations between viral transcript expression patterns, TME subtypes, and patient prognosis.

## Methods

### HPV status prediction classifier

#### Data preparation

Multiple datasets consisting of head and neck cancer samples that were available with HPV status, were used to create the classifier **(Table 1)**.

**Table 1.** Datasets used for HPV status classifier training and testing.

| Cohort ID | Platform | N Samples | Citation | Usage |
| --- | --- | --- | --- | --- |
| GSE65858 | GPL10558 | 269 | Wichmann et al., 2015 [7] | train |
| E-TABM-302 | GPL570 | 73 | Rickman et al., 2008 [8] | train |
| GSE25727 | GPL8432 | 54 | Fountzilas et al., 2012 [9] | train |
| GSE3292 | GPL570 | 1 | Slebos et al., 2006; Chung et al., 2006 [10,11] | train |
| GSE6791 | GPL570 | 42 | Pyeon et al., 2007 [12] | train |
| GSE10300 | GPL570 | 35 | Cohen et al., 2009 [13] | train |
| GSE40774 | GPL13497 | 134 | Keck et al., 2015 [14] | test |
| TCGA_HNSC | RNA-seq | 498 | The Cancer Genome Atlas Network, 2015 [15] | test_size - 100 samples (~20%) with StratifiedShuffleSplit, the rest are train samples |

The data were obtained both using various microarray platforms and using RNA sequencing (RNA-seq). Training and testing were performed with independent datasets and both cohorts contained data from different RNA protocols.

#### TCGA labels preparation

The HPV status of TCGA samples was assigned based on a previous TCGA report [15]. For the samples missing HPV status in the TCGA annotation, Pathseq (https://software.broadinstitute.org/pathseq/) was used as described further in the “Determination of HPV viral status” section in the methods. Pathseq output for the following serotypes was used: Alphapapillomavirus_12, Alphapapillomavirus_4, Alphapapillomavirus_7 (= HPV18), Human_papillomavirus_5, Human_papillomavirus_9, Human_papillomavirus_type_10, Human_papillomavirus_type_16, Human_papillomavirus_1.

#### Feature preparation

Gene expression values were used to train the RNA classifier. To eliminate batch effect without introducing cohort-based batches, we performed rank transformation across samples. As the initial set of features we used 158 genes described by Ankur et al., to be significantly differentially expressed between HPV-positive and HPV-negative HNSCC tumors [16].

#### Classifier training, tuning and testing

As a machine learning model, we implemented the gradient boosting LGBM Classifier. Hyperparameters were chosen using 5-fold cross-validation and F1-weighted metric was chosen for performance assessment. After the training step, the set of hyperparameters that showed the best quality on cross-validation was evaluated to determine the importance of the gene features. The feature with the lowest importance score was removed from the dataset. The subsequent training step was carried out without this gene. Iterations continued until 144 features remained. Finally, the set of features and hyperparameters was chosen that maximized the F1-weighted score. The list of features selected is listed in **Supplementary Table 1**. After this final feature selection step we used one final tuning of hyperparameters. Model testing was performed on a hoold-out independent dataset, which included 20% of the the TCGA HNSCC cohort and another microarray-based cohort (**Table 1**).

### Microarray data processing

Raw and processed microarray data were downloaded from GEO. Expression was re-processed from raw files, if possible, using affygcRMA and oligo R packages. All affymetrix datasets with available CEL files were re-normalized using the gcRMA package with default parameters. Illumina array data were downloaded from GEO as is.

### RNA-seq processing

RNA-seq data was processed from raw reads as described by Bagaev et al. [17]. Briefly, the reads were aligned with Kallisto v0.42.4 and annotated using GENCODEv23 transcripts 69. The expression of 20,,062 protein coding genes was quantified as transcripts per million (TPM) and log2- transformed [18].

### WES processing

Alignment: low quality reads were filtered using FilterByTile/BBMap v37.90 and aligned to human reference genome GRCh38 (GRCh38.d1.vd1 assembly) using BWA v0.7.17. Duplicate reads were removed using Picard’s v2.6.0 MarkDuplicates; indels were realigned by IndelRealigner and recalibrated by BaseRecalibrator and ApplyBQSR (last three tools from GATK v3.8.1).

Variant calling: Both germline and somatic single nucleotide variations (sSNVs), small insertions and deletions were all detected using Strelka v2.9. All variants, insertions and deletions were annotated using Variant Effect Predictor v92.1. Copy number alterations were evaluated with a customized version of Sequenza v2.1.2. Tumor cellularity (purity) estimation was determined as purity estimations (CPE) as previously described [19].

### Tumor microenvironment (TME) classification

We collected three publicly available HNSCC datasets from the GEO and SRA databases (<u>GSE30784</u> [20], <u>GSE40774</u> [14], <u>GSE65858</u> [7], one internal cohort, and the TCGA-HNSC project [15] (total number of samples = 912). Clinical and mutation data were downloaded from the GDC TCGA data portal (MC3 dataset). Transcriptomic data were downloaded from the USCS XENA portal (https://xena.ucsc.edu/) as TPM units. We excluded samples that didn’t pass the quality control due to one of the following reasons: PCA outlier, low correlation with others within the cohort (<0.8 for Affymetrix platforms, < 0.65 for Illumina platforms), low coverage and low phred scores for the RNA-seq, high non-human tissue contamination (>3%), or high percentage of duplicates (>80%). We developed an HPV status prediction algorithm on samples that passed quality control and selected 266 HPV-positive samples from public datasets for further TME subtypes identification. Twenty expression signatures from Bagaev et al. [17] and Batista da Costa et al. [21] were calculated using ssGSEA [22]. The PI3K pathway activity score was calculated using PROGENy [23]. Raw signature scores were median centered and mad scaled within a platform-based batch. TME subtype classification was based on Louvain clustering [24] on scaled signature scores.

### TME validation

To validate the TME classification, two independent HNSCC cohorts from Massachusetts Eye and Ear were utilized (n = 142 and n = 54 RNA-seq samples). Using the developed HPV-status prediction algorithm 108 and 45 HPV-positive samples from these cohorts were selected, respectively. To predict TME subtypes we used the K-nearest neighbor algorithm on pre-selected signatures (**Table 3**) with K=45.

### Determination of HPV viral status, viral expression, subtypes, and host-viral chimeric reads

Viral read identification was based on the GATK Pathseq software kit [25] with quantitative assessment expressed in VRM (viral read per million human reads). Viral status and type verification were performed using the VIRTUS pipeline [26]. The threshold for determining viral status as “positive” was chosen at 2 VRM. At this threshold, the number of raw reads makes it possible to evaluate the expression of viral transcripts. Mapping, score, and quantification of viral transcripts was also analyzed by ViGEN [27] for HPV16 positive (HPV16+) samples (NCBI Reference Sequence: NC_001526). HPV16+ samples were classified into four distinct subtypes (E2/E5, E6/E7, E1/E4, L1/L2) based on unsupervised Louvain clustering [24] of the median scaled TPM-transformed viral gene expression. Additionally, we identified viral-host chimeric reads using Vi-Fi software [28]. The presence of viral-host chimeric reads was interpreted as viral integration in the host genome.

## Results

### Development of single sample host expression-based HPV status predictor

To develop an ML-based algorithm that predicts HPV status, publicly available expression data (n =1,013 HNSCC samples) was used for training (n = 799) and validation (n = 214) **(Figure 1A**, **Table 1)**. The initial set of features for model development was composed of 158 genes distinguishing between HPV-positive and HPV-negative tumors according to literature review [16]. To eliminate the platform batch effect, we included both RNA-seq and array platforms in training and validation sample sets and used rank-transformation of expression values for a selected set of genes. Using the ML-based feature selection algorithm we subsequently reduced the number of features to 144 genes (**Supplementary Table 1**). After the final step of hyperparameter tuning on this feature set the HPV status classifier was validated on an independent set of samples (n = 214) and showed high accuracy in predicting the HPV status for a single sample (ROC-AUC = 0.985) **(Figure 1B)**.

**Figure 1.**
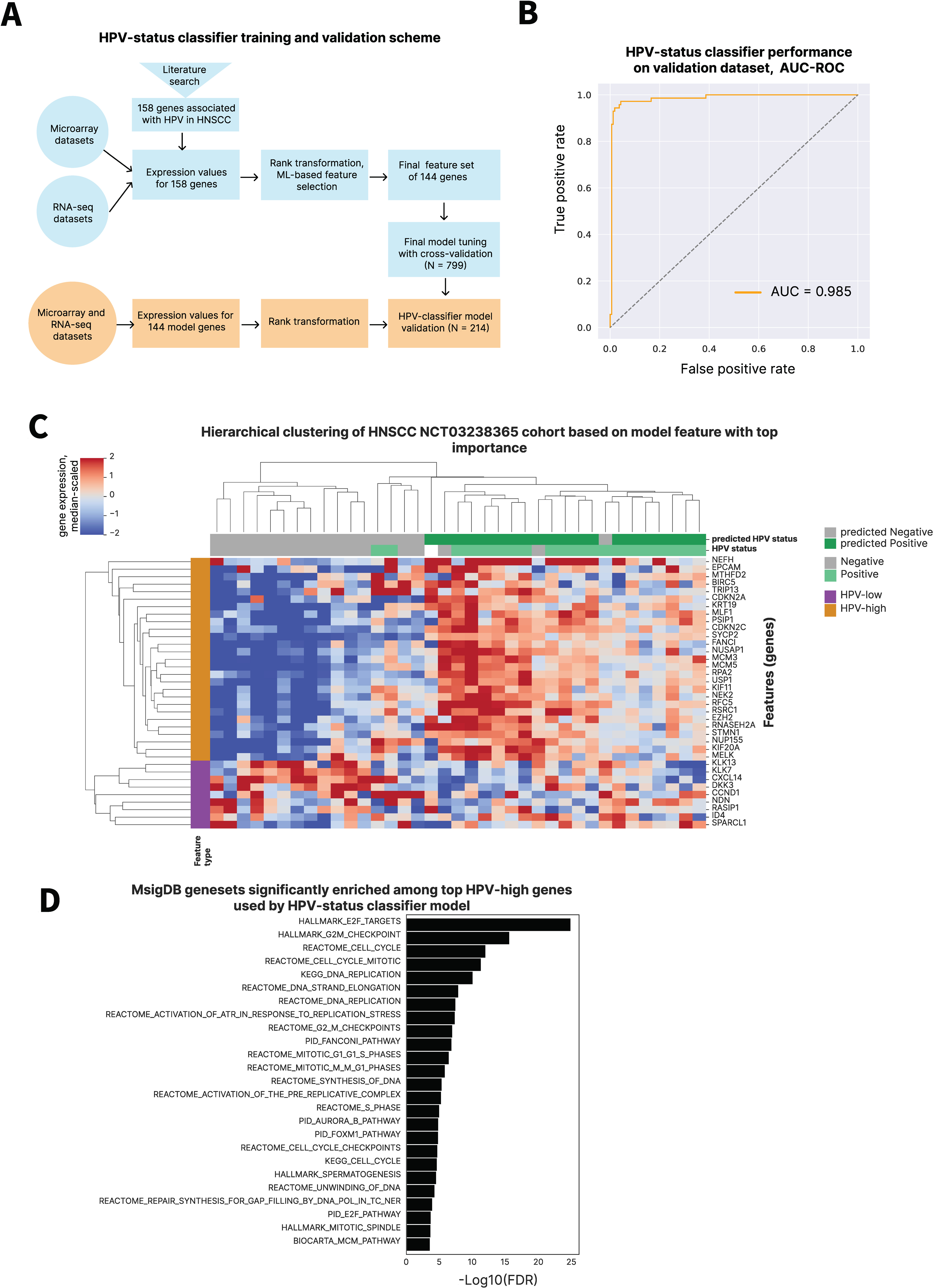
Expression-based ML classifier accurately predicts HPV status. **A.** Expression-based HPV status predictor training and validation procedure. **B.** ROC-AUC for HPV status predictor. **C.** Heatmap showing average linkage clustering of the median scaled expression values for top 25 model genes having the highest feature importance metric for the NCT03238365 cohort (n = 37). **D.** Barplot of the most enriched gene sets among top 16 HPV-high model genes. X-axis shows - Log10(False Discovery Rate) values for each gene set.

In addition, the model was validated using a cohort of HNSCC samples from the clinical trial NCT03238365 [29]. Оnly pre-treatment samples were included to avoid influence of the treatment on the prediction (n = 37). The HPV status classifier achieved a weighted F1-score of 83.4%, correctly predicting the HPV status for 31 of 36 samples. Using this cohort, we demonstrated clear separation of samples into predicted HPV-positive and HPV-negative groups based on expression levels of the top 25 model genes having the highest feature importance metric **(Figure 1C)**. Then, gene set overlap analysis was applied using gene collections v6.1 from MsigDB [30] separately to 16 HPV-high and 9 HPV-low genes (correlating with high or low probability of HPV-driven HNSCC, respectively) **(Figure 1D)**. HPV-high genes were enriched with gene sets of proliferation, mitotic spindle, and cell cycle, and included known genes such as *CDKN2A* and and *CDKN2B*. HPV-low genes did not overlap significantly with any of the gene sets tested.

### HPV-positive HNSCC tumors can be characterized by five distinct tumor microenvironment subtypes

To characterize the TME, our HPV status prediction algorithm was used, and 266 HPV-positive samples from publicly available expression datasets were selected **(Figure 2A)**. We generated 20 Fges representing various immune populations (e.g., Treg cells, B cells, effector cells), stromal components (e.g., angiogenesis, cancer-associated fibroblasts [CAFs]), and tumor properties (e.g., proliferation rate, basal/keratinization). For each of the 266 HPV-positive samples, 19 signature activity scores were calculated using ssGSEA and PI3K pathway activity scores. Further applying unsupervised dense Louvain clustering [24] to these scores revealed 5 subtypes characterized by distinct TME composition and tumor features. Based on enriched signatures they were termed: moderately immune-enriched (IE/M), immune-enriched B cell (IE/B), fibrotic (F), immune-desert (D), immune-enriched luminal-like (IE/L) **(Figure 2B)**. Each subtype was characterized by a distinct signature enrichment pattern. For example, there was a high CAF signature in fibrotic TMEs and a high PI3K pathway score in immune-desert TMEs **(Figure 2C)**. While all three immune-enriched subtypes have relatively higher signature values compared to the other TMEs, samples in the IE/B subtype were enriched in B cell and follicular helper T cell (Tfh) signatures. Further, samples in the IE/L subtype were distinguished from other inflamed tumors by a very low basal/keratinization signature (**Figure 2C)**. There were no associations between the tumor site and the TME subtype **(Figure S1A)**. Importantly, TME subtypes were associated with OS and patient prognosis. Tumors with an immune-enriched microenvironment showed the highest survival rates, whereas patients with a fibrotic TME subtype had poor survival **(Figure 2D)**. Independent validation of the TME subtypes was performed on an internal HPV-positive cohort (n = 45) **(Figure 2E, S1B)**.

**Figure 2.**
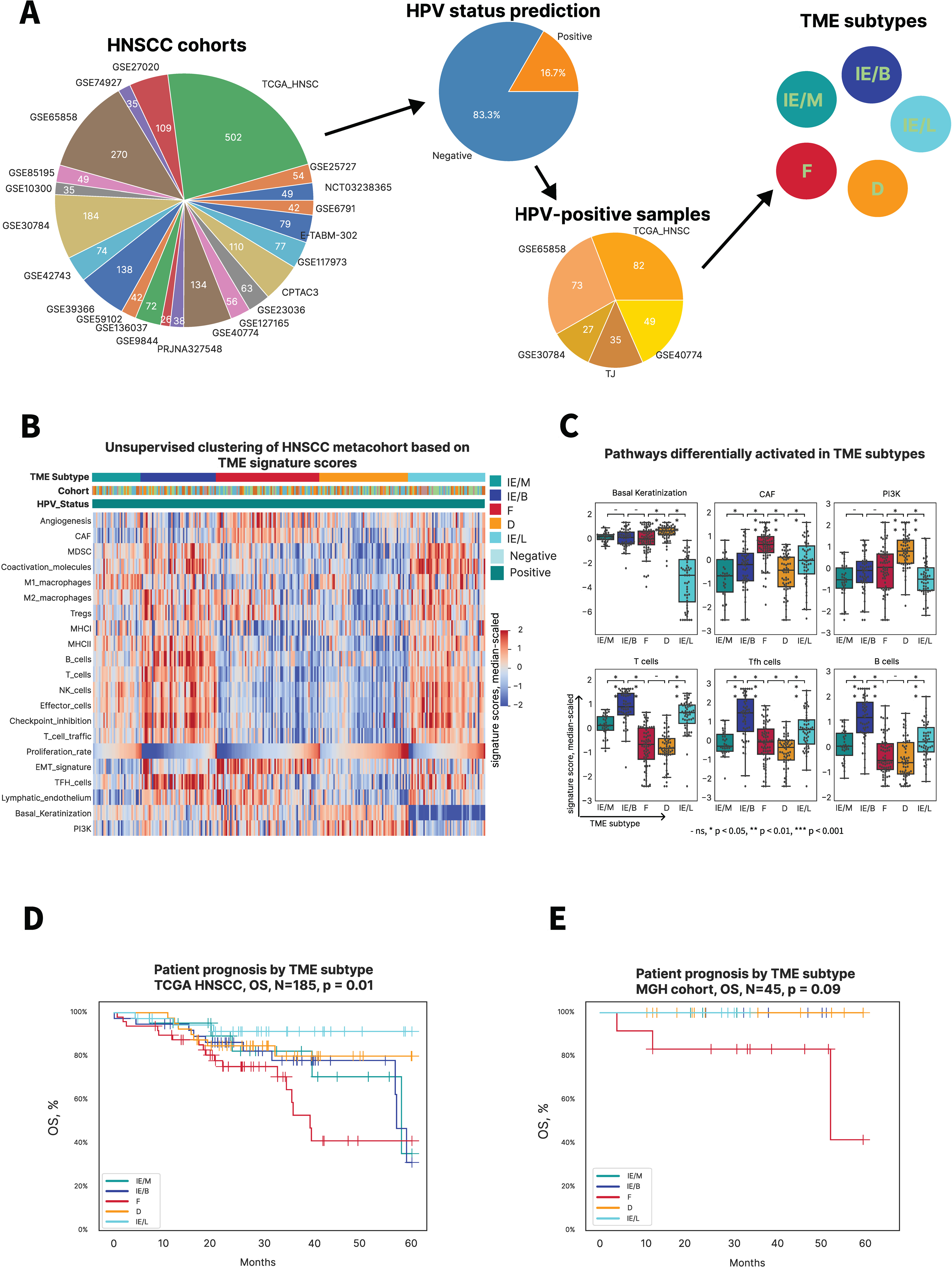
A novel tumor microenvironment classification in HPV-positive HNSCC predicts survival outcome. **A.** HNSCC expression datasets collection followed by HPV status prediction. Gene expression signature activation scores of HPV-positive samples (N = 266) were then used for dense clustering which revealed 5 distinct TME clusters. **B.** Heatmap showing functional gene expression signature activation scores for a meta-cohort of HPV-positive samples. The X axis represents samples, and the Y axis shows different signatures. **C.** Boxplots showing statistically significant differences in signature scores among TME subtypes. - ns, * p < 0.05, ** p < 0.01, *** p < 0.001. **D.** Overall survival (OS) of patients from the TCGA HNSCC HPV-positive cohort stratified by TME subtype. **E.** OS of patients from the HNSCC HPV-positive validation cohort stratified by TME subtype (Supplement figure 1C).

### HPV transcript expression stratifies HPV-positive HNSCCs into four subtypes associated with prognosis

To further elucidate the specific factors associated with survival of HPV-positive HNSCC patients, patterns of HPV-specific transcript expression were investigated. By retrieving viral transcript expression data from bulk RNA-seq and using unsupervised clustering of viral transcript expression values in the TCGA HNSCC HPV-positive dataset, 4 HPV-related subtypes were identified. Each subtype was enriched for distinct viral transcripts expressed at different life cycle stages: E2/E5, E6/E7, E1/E4 and L1/L2 [31] **(Figure 3A)**. Survival analysis of these subtypes and HPV-negative samples showed that the E6/E7 cluster was associated with the worst OS among HPV-positive samples, approaching HPV-negative tumors. The best prognosis was in patients belonging to the E2/E5 viral expression subtype **(Figure 3B)**. To further investigate patterns of viral gene expression in HPV-positive squamous cell carcinoma (SCC), we performed the same clustering analysis using the TCGA cervical SCC (CESC) dataset and discovered the same 4 viral subtypes as for HNSCC **(Figure 3C).** Patients with the E6/E7 viral subtype of CESC had statistically indistinguishable progression-free survival (PFS) from HPV-negative CESC samples and the poorest prognosis among all HPV-positive subtypes **(Figure 3D)**. Prognostic stratification of all viral subtypes in CESC closely resembled that for HNSCC with the E2/E5 cluster associated with the best prognosis (OS in HNSCC and PFS in CESC). Independent validation of HPV subtypes was performed on an internal HPV-positive cohort (n = 108) and revealed similar viral subtypes **(Figure S1C, S1D)**.

**Figure 3.**
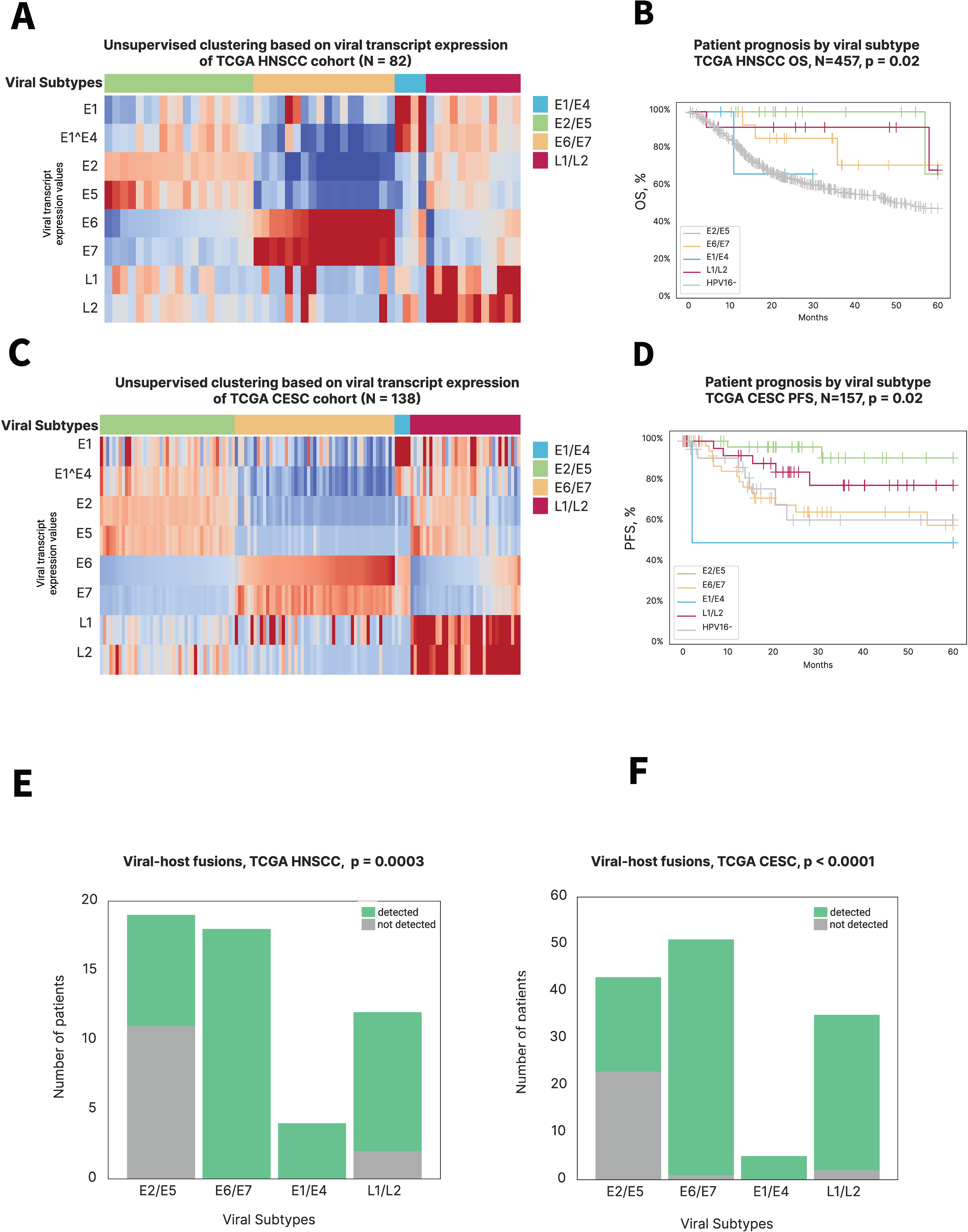
Four viral subtypes based on HPV transcript expression in HNSC and CESC are associated with survival and viral genome integration. **A. and C.** Heatmaps showing HPV16 transcript expression scores among distinct viral subtypes in TCGA HNSC and TCGA CESC datasets respectively. The X axis represents samples, and the Y axis shows HPV16 genes. **B.** Overall survival of patients from TCGA HNSC cohort stratified by viral subtypes classification. **C.** Progression free survival of patients from TCGA CESC cohort stratified by viral subtypes classification. **D. and F.** Amount of samples with or without detected chimeric viral-host reads per viral subtypes among TCGA HNSC HPV-positive and TCGA CESC HPV-positive cohorts respectively.

Detection of viral-host mRNA fusions showed that the E2/E5 subtype was enriched for samples without HPV-genome integration, suggesting that HPV episomal DNA status and an E2/E5 expression pattern may drive an inflamed microenvironment and improved prognosis **(Figure 3E)**. These findings were validated on CESC TCGA samples **(Figure 3F).**

### E2/E5 HPV subtype associates with immune-enriched subtypes

Utilizing both viral transcript and TME subtypes of the TCGA-HNSCC cohort, we found that the E2/E5 HPV subtype was associated with an immune-enriched TME (74%) **(Figure 4A, S1E).** This finding was in concordance with higher OS of E2/E5 and immune-enriched samples (IE/M, IE/B, IE/L) compared to the other subtypes. On the contrary, the E6/E7 subtype was associated with an immune-desert TME (50%) and comprised more samples with fibrotic TMEs (28%) than any other viral subtype. Among all HPV clusters, the E6/E7 subtype had the lowest levels of T and B cell gene expression signatures, resembling those of HPV-negative HNSCCs **(Figure 4B)**. Patients with such tumors had poor prognosis according to both classifications. Interestingly, the E6/E7 subtype did not include any IE/L samples, suggesting that these HNSCCs were less differentiated and more resembled “basal” keratinocytes, which could explain the aggressiveness of such tumors. On the other hand, samples with the lowest basal signature from the IE/L subtype were mostly clustered within the L1/L2 viral subtype, consistent with the fact that L1 and L2 capsid proteins are more highly expressed toward the layers of squamous epithelium [31]. Taking into account higher differentiation of such keratinocytes, this finding is also in accordance with the good prognosis that was observed for patients from the IE/L and L1/L2 subtypes.

**Figure 4.**
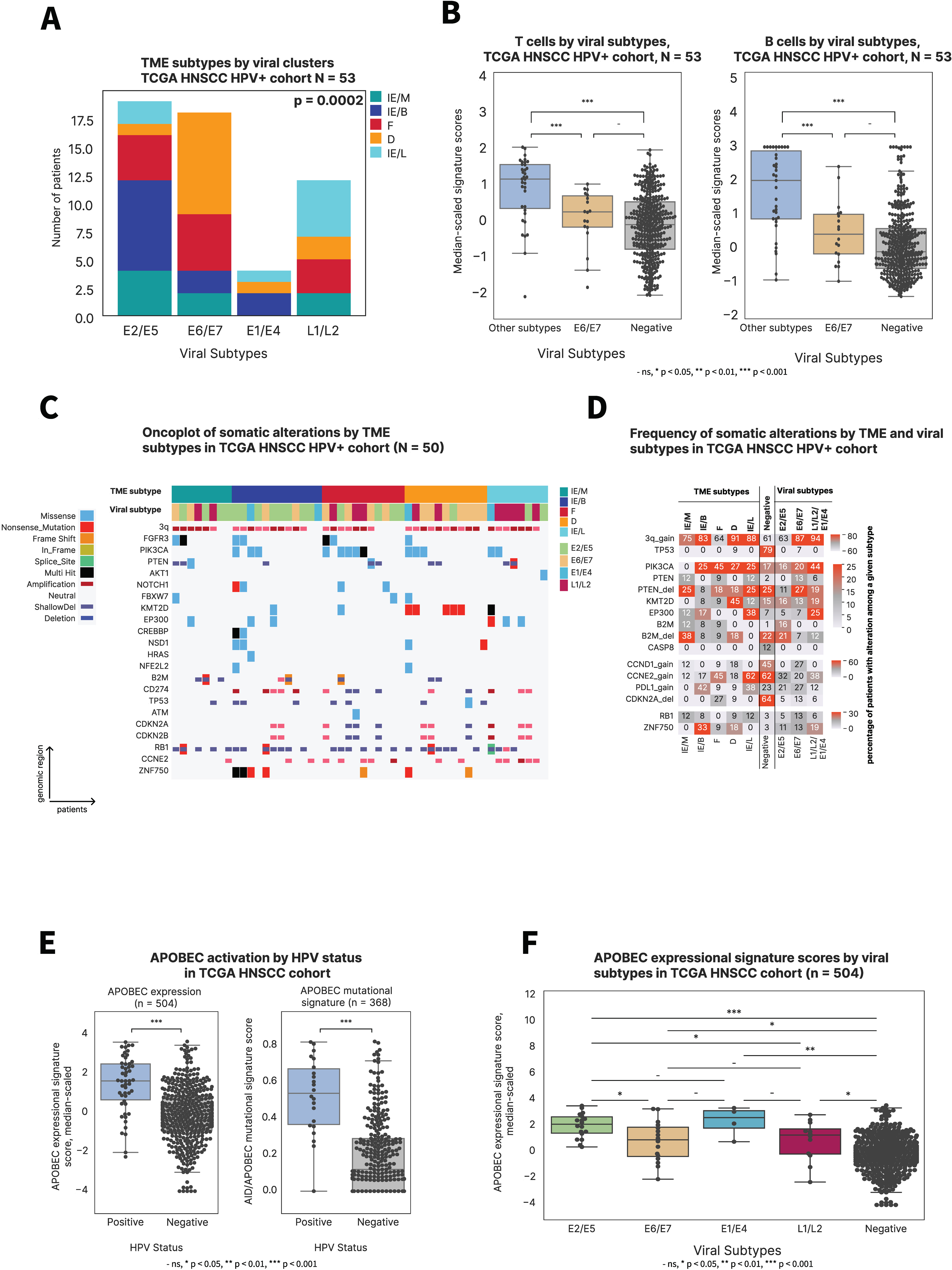
TME subtypes are associated with viral classification and tumor genetics. **A.** Distribution of TME subtypes in each viral subtypes in TCGA HNSC HPV-positive cohort. **B.** Boxplots showing differences in B- and T-cells signature expression scores across all TCGA HNSC samples combined into three groups: E6/E7 subtype (n = 18), other HPV subtypes (n = 35; E2/E5, E1/E4, L1/L2 together) and HPV-negative (n = 388). **C.** Oncoprint and **D.** alteration rate of driver somatic alterations among different TME and viral subtypes for TCGA HNSCC cohort. **D.** Boxplots showing APOBEC genes expression signature scores and mutational signature scores in HPV-positive and HPV-negative HNSCCs. **E.** Boxplots showing APOBEC genes expression signature scores in among viral subtypes of HPV-positive and HPV-negative HNSCCs.

### Distinct genetic features of various viral and TME subtypes of HPV-positive HNSCCs

Major differences in genomic landscapes of HPV-positive and HPV-negative HNSCCs have been characterized previously and were confirmed in this study, including lower rates of CDKN2A(p16) deletions and somatic mutations in genes *TP53*, *MYC*, *CCND1*, *FAT1*, *EGFR*, and *CASP8* and higher rates of *GSK3B*, *NBPF1,* and *TRAF3* mutations in HPV-positive tumors **(Figure S2A, S2B)**. Interestingly, we revealed a statistically significant difference in *PIK3CA* hotspot mutations among HPV-positive and HPV-negative tumors **(Figure S2C)** that was previously suspected but “data remained insufficient to establish a pattern” [32]. In this study, among all somatic mutations in the *PIK3CA* gene, the alterations *E542K*, *E545K* and *E546K* were found to be more prevalent in HPV-positive HNSCCs (p = 0.0002, n(HPV+) = 50, n(HPV-) = 391) **(Figure S2D)**. Further, the rates of prevalent driver genomic alterations among TME and viral subtypes were compared were compared. We observed that tumors characterized by E6/E7 HPV and D and F TME subtypes were genetically more similar to HPV-negative tumors subtypes **(Figure 4C, 4D).** For example, they more frequently harbored p16 deletions. Interestingly, these samples also had higher rates of *CCND1* copy number gains, similar to HPV-negative tumors and consistent with CCND1 higher expression in HPV-negative tumors **(Figure 1C).** On the contrary, immune-enriched subtypes and subtypes that were associated with E2/E5 showed high rates of B2M alterations, both mutations and deletions, suggesting these tumors might rely on such alterations as a mechanism of immune escape. Another highly noticeable alteration associated with immune escape - PDL1 copy number gain is observed in high rates among most immune-enriched TME subtypes - IE/B and IE/L **(Figure 4C, 4D)**. Finally, the immune-desert TME subtype was significantly enriched with *KMT2D* truncating mutations, whereas other subtypes harbored lower rates and mostly missense mutations in this chromatin-remodeling gene **(Figure 4C).**

### APOBEC activation in HPV-positive HNSCC

APOBEC deaminases have been shown to play an important role in the mutagenesis of HPV- positive HNSCC [6,33,34]. Here, we confirmed that APOBEC activation was higher in HPV- positive compared to HPV-negative samples both by mutational signature and gene expression signature analysis **(Figure 4E).** Among viral subtypes, APOBEC expression was higher in E2/E5 tumors **(Figure 4F**), consistent with theories that APOBEC is upregulated in response to episomal HPV as a host defense mechanism. However, in the TCGA CESC cohort a statistically significant difference was observed only when comparing APOBEC expression in HPV-positive and HPV-negative **(Figure S2E**), but not among viral subtypes and not at the AID/APOBEC mutational signature level **(Figure S2E, S2F).**

## Discussion

HNSCC is the sixth most common cancer worldwide and is comprised of two distinct subtypes: carcinogen-driven and viral-driven. Although HPV-positive HNSCC has unique clinical and molecular characteristics from HPV-negative HNSCC, further efforts are needed to identify patient subgroups for personalized treatment. Here, we leveraged an AI-driven algorithm to understand the tumor, TME, and contributions of HPV to tumor progression, discerning an intriguing relationship between viral transcript expression, TME subtypes, and prognosis in HPV-associated HNSCCs. The TME subtypes that we identified - immune-enriched, highly immune and B-cell enriched, fibrotic, immune-desert, and immune-enriched luminal - exhibit distinct correlations with survival and prognosis, highlighting the prognostic implications of the TME composition. Interestingly, the HPV E2/E5 transcript subtype, which was associated with an immune-enriched TME and an improved prognosis, was also enriched in tumors without HPV-genome integration. In contrast, the E6/E7 subtype was associated with a fibrotic or desert TMEs, characterized by reduced T-cell and B-cell gene expression and poorer survival. Therefore, the E6/E7 subtype might represent a more immune-resistant phenotype, which could necessitate treatment escalation or selection of non-immune-mediating therapies.

These findings shed light on the intricate interplay between viral oncogenes, host immune response, and clinical outcomes in HPV-associated HNSCC and underscore the potential utility of viral transcript profiling and TME characterization in prognostication and guiding personalized therapy. Despite the extensive and variable datasets used in this analysis, it is important to note that further validation is necessary to confirm these associations and assess their potential implications in the clinical setting. Moreover, exploring the mechanistic basis of these observations could uncover novel insights into the pathogenesis of HPV-associated HNSCC and pave the way for new therapeutic approaches.

Our study emphasizes the importance of adopting a multi-dimensional approach to understanding tumor biology, which integrates viral genomics, host immunology, and clinical data to optimize patient care in HPV-associated HNSCC.

## Declarations

### Ethics approval and consent to participate

Not applicable.

### Consent for publication

Not applicable.

### Availability of data and material

The data and code will be deposited online and publicly available at the time of publication.

### Competing interests

LW has received research funding from Lilly and Novartis. DF has received research funding from BMS, Calico, Predicine, BostonGene, and Neogenomics. DF owns stocks from Illumina, Roche, and Qiagen.

All BostonGene authors were employees thereof at the time the study was performed.

### Funding

This research was funded by BostonGene, Corp.

### Authors’ contributions

Anastasia Nikitina - conceptualization, results interpretation, original draft writing and revising the manuscript

Daria Kiriy - TME part analysis: pipelines launching, processing raw data, computing TME clusters, validation; WES/RNAseq analysis; figures construction; methods section writing. Andrey Tyshevich - viral part analysis: pipelines launching, processing raw data, computing viral clusters and integrations

Z. Antysheva - HPV status predictor training and validation

D. Tychinin - viral part analysis, methods section writing.

Anastasia Sobol - figures construction and methods description for HPV status predictor

Vladimir Kushnarev - supervision and discussion

Nara Shin - supervision, discussion, and revising the manuscript

Jessica H. Brown - discussion and revising the manuscript

Krystle Kuhs - validation cohorts preparation, reviewing the manuscript

James Lewis Jr. - validation cohorts preparation, reviewing the manuscript

Robert Ferris - validation cohorts preparation, reviewing the manuscript

Lori Wirth - Conceptualization, supervision, reviewing the manuscript

Nikita Kotlov - Conceptualization, supervision, reviewing the manuscript

Daniel L. Faden - conceptualization, results interpretation, validation cohorts preparation, and revising the manuscript

All authors read and approved the final manuscript.

## Data Availability

All data produced in the present work are contained in the manuscript or available via the referenced works.

## Acknowledgements

The results published here are in whole or part based upon data generated by The Cancer Genome Atlas (TCGA) managed by the NCI and NHGRI. Information about TCGA can be found at http://cancergenome.nih.gov/.

## Supplemental figure legends

**Figure S1. Validation of TME subtypes**

**A.** Distribution of TME subtypes by tumor site.

**B.** Heatmap showing functional gene expression signature activation scores for an internal cohort of HPV-positive samples (n=45). The X axis represents samples, and the Y axis shows different signatures.

**C.** Heatmap showing HPV16 transcript expression scores among distinct viral subtypes in an internal cohort (n=108). The X axis represents samples, and the Y axis shows HPV16 genes.

**D.** Overall survival of patients from an internal cohort (n=108) stratified by viral subtypes classification.

**E.** Percentage of TME subtypes in each viral subtypes in TCGA HNSC HPV-positive cohort (n=53).

**Figure S2. Differential genomic events in HPV+ vs HPV- HNSCCs.**

**A.** Oncoprint and **B.** alteration rate of differentially present somatic alterations in HPV-positive and HPV-negative HNSCCs

**B.** Oncoprint and **D.** alteration rate of hotspot somatic mutations in PIK3CA gene in HPV- positive and HPV-negative HNSCCs

**E.** Boxplots showing APOBEC genes expression signature scores and mutational signature scores in HPV-positive and HPV-negative HNSCCs in TCGA CESC cohort.

**F.** Boxplots showing APOBEC genes expression signature scores in among viral subtypes of HPV-positive and HPV-negative HNSCCs in TCGA CESC cohort (n=293).

## References

1. Fridman WH, Pagès F, Sautès-Fridman C, Galon J. The immune contexture in human tumours: impact on clinical outcome. Nat Rev Cancer. 2012;12:298–306.

2. Chen DS, Mellman I. Oncology Meets Immunology: The Cancer-Immunity Cycle. Immunity. 2013;39:1–10.

3. Liu T, Han C, Wang S, Fang P, Ma Z, Xu L, et al. Cancer-associated fibroblasts: an emerging target of anti-cancer immunotherapy. J Hematol Oncol. 2019;12:86.

4. Schaaf MB, Garg AD, Agostinis P. Defining the role of the tumor vasculature in antitumor immunity and immunotherapy. Cell Death Dis. 2018;9:115.

5. Lechien JR, Descamps G, Seminerio I, Furgiuele S, Dequanter D, Mouawad F, et al. HPV Involvement in the Tumor Microenvironment and Immune Treatment in Head and Neck Squamous Cell Carcinomas. Cancers (Basel). 2020;12:1060.

6. Faden DL, Ding F, Lin Y, Zhai S, Kuo F, Chan TA, et al. APOBEC mutagenesis is tightly linked to the immune landscape and immunotherapy biomarkers in head and neck squamous cell carcinoma. Oral Oncol. 2019;96:140–7.

7. Chakravarthy A, Henderson S, Thirdborough SM, Ottensmeier CH, Su X, Lechner M, et al. Human Papillomavirus Drives Tumor Development Throughout the Head and Neck: Improved Prognosis Is Associated With an Immune Response Largely Restricted to the Oropharynx. J Clin Oncol. 2016;34:4132–41.

8. Bagaev A, Kotlov N, Nomie K, Svekolkin V, Gafurov A, Isaeva O, et al. Conserved pan-cancer microenvironment subtypes predict response to immunotherapy. Cancer Cell. 2021;39:845–865.e7.

9. Goldman MJ, Craft B, Hastie M, Repečka K, McDade F, Kamath A, et al. Visualizing and interpreting cancer genomics data via the Xena platform. Nat Biotechnol. 2020;38:675–8.

10. Aran D, Sirota M, Butte AJ. Systematic pan-cancer analysis of tumour purity. Nat Commun. 2015;6:8971.

11. Chen C, Méndez E, Houck J, Fan W, Lohavanichbutr P, Doody D, et al. Gene expression profiling identifies genes predictive of oral squamous cell carcinoma. Cancer Epidemiol Biomarkers Prev. 2008;17:2152–62.

12. Keck MK, Zuo Z, Khattri A, Stricker TP, Brown CD, Imanguli M, et al. Integrative Analysis of Head and Neck Cancer Identifies Two Biologically Distinct HPV and Three Non-HPV Subtypes. Clinical Cancer Research. 2015;21:870–81.

13. Wichmann G, Rosolowski M, Krohn K, Kreuz M, Boehm A, Reiche A, et al. The role of HPV RNA transcription, immune response-related gene expression and disruptive TP53 mutations in diagnostic and prognostic profiling of head and neck cancer. Int J Cancer. 2015;137:2846–57.

14. Batista Da Costa J, Gibb EA, Bivalacqua TJ, Liu Y, Oo HZ, Miyamoto DT, et al. Molecular Characterization of Neuroendocrine-like Bladder Cancer. Clinical Cancer Research. 2019;25:3908–20.

15. Yi M, Nissley DV, McCormick F, Stephens RM. ssGSEA score-based Ras dependency indexes derived from gene expression data reveal potential Ras addiction mechanisms with possible clinical implications. Sci Rep. 2020;10:10258.

16. Schubert M, Klinger B, Klünemann M, Sieber A, Uhlitz F, Sauer S, et al. Perturbation-response genes reveal signaling footprints in cancer gene expression. Nat Commun. 2018;9:20.

17. Blondel VD, Guillaume J-L, Lambiotte R, Lefebvre E. Fast unfolding of communities in large networks. 2008.

18. Yasumizu Y, Hara A, Sakaguchi S, Ohkura N. VIRTUS: a pipeline for comprehensive virus analysis from conventional RNA-seq data. Bioinformatics. 2021;37:1465–7.

19. Bhuvaneshwar K, Song L, Madhavan S, Gusev Y. viGEN: An Open Source Pipeline for the Detection and Quantification of Viral RNA in Human Tumors. Front Microbiol. 2018;9:1172.

20. Nguyen N-PD, Deshpande V, Luebeck J, Mischel PS, Bafna V. ViFi: accurate detection of viral integration and mRNA fusion reveals indiscriminate and unregulated transcription in proximal genomic regions in cervical cancer. Nucleic Acids Res. 2018;46:3309–25.

21. Liberzon A, Birger C, Thorvaldsdóttir H, Ghandi M, Mesirov JP, Tamayo P. The Molecular Signatures Database (MSigDB) hallmark gene set collection. Cell Syst. 2015;1:417–25.

22. Burley M, Roberts S, Parish JL. Epigenetic regulation of human papillomavirus transcription in the productive virus life cycle. Semin Immunopathol. 2020;42:159–71.

23. Faden DL, Thomas S, Cantalupo PG, Agrawal N, Myers J, DeRisi J. Multi-modality analysis supports APOBEC as a major source of mutations in head and neck squamous cell carcinoma. Oral Oncol. 2017;74:8–14.

24. Faden DL, Kuhs KAL, Lin M, Langenbucher A, Pinheiro M, Yeager M, et al. APOBEC Mutagenesis Is Concordant between Tumor and Viral Genomes in HPV-Positive Head and Neck Squamous Cell Carcinoma. Viruses. 2021;13:1666.

